# Clinical features of coronavirus disease 2019 (COVID-19) in a cohort of patients with disability due to spinal cord injury

**DOI:** 10.1101/2020.04.20.20072918

**Authors:** Miguel Rodríguez-Cola, Irena Jiménez-Velasco, Francisco Gutiérrez-Henares, Elisa López-Dolado, Claudia Gambarrutta-Malfatti, Eduardo Vargas-Baquero, Ángel Gil-Agudo

**Author notes:** ***Corresponding author*** Ángel Gil-Agudo, MD PhD, Head of Department of Rehabilitation, Hospital Nacional de Parapléjicos of Toledo (Spain). Address: Hospital Nacional de Parapléjicos. Finca La Peraleda s/n. 45071 Toledo (Spain).

## Abstract

**Study design:** Cohort study of patients with spinal cord injury (SCI)

**Objectives:** To describe the clinical and analytical features of a Covid-19 infected cohort with SCI to contribute new knowledge for a more accurate diagnosis and to outline prevention measures.

**Setting:** This study was conducted at the National Hospital for Paraplegics (Toledo, Spain).

**Methods:** A cohort analysis of seven patients with SCI infected by Covid-19 was carried out. Diagnosis was confirmed with reverse transcriptase polymerase chain reaction (RT-PCR) of nasal exudate or sputum samples. Clinical, analytical and radiographic findings were registered.

**Results:** RT-PCR detected COVID-19 infection in all patients, affecting males and people with a cervical level of injury more often (5 out 7). The average delay for diagnostic confirmation was 4 days (interquartile range, 1-10). Fever was the most frequent symptom (6 out of 7). The second most common symptom was asthenia (4 out of 7), followed by dyspnea, cough and expectoration (3 out of 7 for each symptom). The MEWS score for Covid-19 severity rating was classified as severe in 5 out of 7. All but one patient showed radiological alterations evident in chest X-Rays at the time of diagnosis. All patients recovered gradually.

**Conclusion:** Our patients with SCI and Covid-19 infection exhibited fewer symptoms than the general population. Furthermore, they presented similar or greater clinical severity. The clinical evolution was not as pronounced as had been expected. This study recommends close supervision of the SCI population to detect early compatible signs and symptoms of Covid-19 infection.

## Introduction

In late December 2019, an epidemic of severe acute respiratory syndrome coronavirus 2 (SARS-CoV-2) began in Wuhan^1^ and spread rapidly throughout the world. The World Health Organization (WHO) declared the 2019 coronavirus disease (Covid-19) pandemic on March 11, 2020 and as of April 2, 2020, a total of 1,002,159 cases were confirmed worldwide with 51,485 deaths^2^. The first confirmed imported case of Covid-19 was detected in Spain in La Gomera, (Canary Islands) on January 31, 2020^3^. Since then and up to April 2, 2020, a total of 110,286 cases have been confirmed in Spain, including 10,096 deaths^2^. On March 17, 2020, the first case of Covid-19 was confirmed at the Hospital Nacional de Parapléjicos (HNP) in Toledo Spain, a 212-bed national spinal cord injury (SCI) monographic hospital classified as a long-term healthcare facility dedicated to the comprehensive rehabilitation of acute SCI as well as people with chronic SCI with multiple comorbidities. The source of the first patient’s infection was close contact with their relative, who was also later diagnosed with Covid-19. Since then, despite having isolated the infected patient in the hospital and used personal protective equipment (PPE) against infection by both the healthcare workers and the non-infected patients’ in quarantine, it was inevitable that new cases of infection were also detected among the patients with up to a total of seven currently confirmed as of April 7, 2020.

SCI involves multiple neurological and systemic complications which drastically interfere with the patient’s quality of life and their life expectation^4^. Several demographic and clinical factors associated with the more aggressive Covid-19 phenotype have been identified: male gender, age over 60 years and the presence of previous comorbidities such as hypertension, diabetes mellitus, obesity, cardiac ischemic disease, lung disease and immunosuppression^5^. It is reasonable to assume SCI as a high-risk comorbidity, mostly due to the presence of respiratory failure as a result of thoracoabdominal muscle weakness^10^ and also because of systemic immunosuppression due to injury^6,7^. Suppresion of the immune system after SCI is due to noradrenergic overactivation and excess glucocorticoid release via hypothalamus-pituitary-adrenal axis stimulation^8^ and also to autoimmunity^9^. Both phenomena, respiratory failure and injury-induced immunosuppression are more pronounced after cervical or high-thoracic compared with low-thoracic SCI levels, which explains why pneumonia is the leading cause of mortality in SCI patients^11^. As Covid-19 is a novel viral respiratory infection, its epidemiology and clinical course among people with SCI is yet unknown. However, recently the first survey has been published which addresses COVID-19-related practices and adaptations among health care professionals working with individuals with SCI^12^. So far, only one case of Covid 19 infection with SCI has been reported^13^. Considering all the secondary complications associated with SCI, it is reasonable to expect a harsh prognosis with Covid-19 infection.

In the present study, the authors aimed to describe the clinical features of the cohort of hospitalized SCI cases with confirmed Covid-19 infection in a monographic hospital in Spain, information which is especially important for a more accurate diagnosis and to prevent future outbreaks.

## Methods

### Participants and inclusion/exclusion criteria

An observational prospective analysis was made of a SCI patient cohort with confirmed Covid-19 infection from March 20 up to April 4, 2020, all of whom had been previously admitted for clinical care at the national SCI monographic hospital in Spain. All of the patients had attended an inpatient treatment regime during the present SARS-CoV2 pandemic. Those patients in whom this infection could not be confirmed were excluded. All participants provided their informed consent. The guidelines of the declaration of Helsinki were followed in every case and the study design was approved by the local Ethics Committee (Hospital Virgen de la Salud, Toledo, Spain, resolution number 504).

The following demographic variables were recorded in the cohort: age, sex, SCI aetiology, neurological level and severity, with the last two variables assessed in accordance with the International Standards for Neurological Classification of SCI^14^.

### Data sources

The main sources of data were the electronic medical records and clinical reports of each patient. Data were recorded of the history of recent physical contacts made between the confirmed case, the diagnostic time delay until RT-PCR confirmation, symptom and signs assessed at onset and evolution, and the laboratory and chest X-rays. In order to avoid biases and to ensure the patients’ data confidentiality, all documents were handled after removing personal or identifying data.

### Variables and definitions

Covid-19 diagnostic confirmation was defined as a positive result detected using the polymerase chain reaction with reverse transcriptase (RT-PCR) of nasal exudate or sputum samples according to the Spanish Ministry of Health and the Castilla La Mancha Government action procedures for cases of Infection of the new coronavirus (SARS-CoV-2)^3^. A *recent physical contact exposure* was defined as a close contact with a family member, caregiver or healthcare professional diagnosed with Covid-19.

Fever was defined as a temperature higher than 37.5ºC. Lymphocytopenia was defined as a lymphocyte count of less than 1500 cells per cubic millimeter (mm^3^). Thrombocytopenia was defined as a platelet count of less than 150.000/ mm^3^. Pneumonia was defined based on the radiological report by a hospital radiologist, classified either as normal, with consolidation in only one lobe, bilateral multilobular, or with a ground glass pattern. In addition, the presence of atelectasis and or pleural effusion was recorded^15^. Laboratory tests included complete chemical analysis including liver and kidney function assessment, electrolytes, C-reactive protein (PCR), lactate dehydrogenase (LDH), ferritin, and blood count.

The Covid-19 severity was defined using the Modified Early Warning System (MEWS)^16^. A non-severe Covid-19 case was defined by a MEWS score ≤ 2. A severe Covid-19 case was defined by a MEWS score > 2 but <5. A critical Covid-19 case was defined by a MEWS score > 5. In the case of pneumonia, we included the CURB-65 score^17^.

### Statistical analysis

Data were expressed as mean or median +/- standard deviation as well as a percentage. As it is a case series with only 7 patients included, further statistical analysis was not performed.

## Results

As of April 4, 2020, 7 cases of Covid-19 have been confirmed among patients with SCI admitted to the HNP. In two cases with an incubation period of 4 and 9 days respectively, the source of infection was a close contact with their relatives who were also subsequently confirmed with Covid-19. In the other five cases, no clear source of infection was identified.

### Demographics

The clinical profile of the cohort is shown in Table 1. The majority were male (5 out of 7; 71.4%), with an average age of 68 years (interquartile range, 34 to 75). A total of 5 out of 7 patients (71.4%) of the cohort presented with a cervical SCI that was sensorimotor complete AIS A in 4 out of 7 of cases (57.1%). A subacute SCI was present in more than half of the patients (4 out of 7), with less than 3 months time from injury. With respect to the SCI aetiology, 4 of the cases were traumatic, 2 of them were vascular and 1 of them, iatrogenic. A total of 4 of the cases had a tracheostomy, and 2 of them required frequent aspirations to remove respiratory secretions before Covid-19 infection. Four of patients had a history of risk factors, such as hypertension (4 out of 7; 57.1%), dyslipidemia (4 out of 7; 57.1%), obesity (3 out of 7; 42.9%) and diabetes mellitus (1 out of 7; 14.3%). Finally, 3 of the patients (42.9%) were either current or previous smokers and lung disease had been previously described.

**TABLE 1.** Clinical features of Covid-19 SCI cohort.

| **TABLE 1. Clinical features of Covid-19 SCI cohort** |  |
| --- | --- |
| **Variable** | **Sample (N=7)** |
| Age– median (IQR) yr | 68 (34-75) |
| Male sex – no./total no. (%) | 5/7 (71.4) |
| **SCI level** |  |
| Cervical– (number/sample (%)) | 5/7 (71.4) |
| Thoracic T1-T6 (number/sample (%)) | 1/7 (14.3) |
| Thoracic T7-T12 (number/sample (%)) | 1/7 (14.3) |
| Lumbosacral (number/sample (%)) | 0/7 (0) |
| **SCI etiology** |  |
| Traumatic (number/sample (%)) | 4/7 (57.1) |
| Vascular– (number/sample (%)) | 2/7 (28.6) |
| Iatrogenic (number/sample (%)) | 1/7 (14.3) |
| **AIS Impairment Scale** |  |
| A (number/sample (%)) | 4/7 (57.1) |
| B (number/sample (%)) | 0/7 (0) |
| C (number/sample (%)) | 2/7 (28.6) |
| D (number/sample (%)) | 1/7 (14.3) |
| E (number/sample (%)) | 0/7 (0) |
| **Tracheostomy** (number/sample (%)) | 4/7 (57.1) |
| Need to remove respiratory secretions(number/sample (%)) | 2/4 (50) |
| **Contact with Covid-19 patient**(number/sample (%)) | 2/7 (28.6) |
| **Incubation period**- media (IQR) dias | 6.5 (4-9) |
| History of Smoking(current or former) (number/sample (%)) | 3/7 (42.9) |
| **Coexisting disorders** (number/sample (%)) |  |
| Any | 0 (0) |
| Hipertension | 4 (57.1) |
| Diabetes | 1 (14.3) |
| Dyslipidemia | 4 (57.1) |
| Obesity | 3 (42.9) |
| Cardiovascular disease | 3 (42.9) |
| Pulmonary disease | 3 (42.9) |
| Chronic renal disease | 0 (0) |
| Cancer | 1 (14.3) |
| **Symptoms of Covid-19 onset (number/sample (%))** |  |
| Fever | 6 (85.7) |
| Dyspnea | 3 (42.9) |
| Cough | 3 (42.9) |
| Fatigue | 4 (57.1) |
| Sputum production | 3 (42.9) |
| Confusion | 2 (28.6) |
| Gastrointestinal symptoms | 1 (14.3) |
| **ModifiedEarlyWarningSystem (MEWS)** |  |
| 0-2 points(number/sample (%)) | 2/7 (28.6) |
| > 3 points (number/sample (%)) | 5/7 (71.4) |
| **CURB-65 – if pneumonia** |  |
| 0-1 points (number/sample (%)) | 1/5 (20) |
| > 1 points (number/sample (%)) | 4/5 (80) |

### Covid-19 clinical features

The median diagnostic time delay, defined as the time period from the onset of symptoms to confirmed Covid-19 infection with RT-PCR, was 4 days (interquartile range, 1-10), and was less than 6 days in 6 out of 7 (85.7%) of cases. Fever was presented in 6 out of 7 (85.7%) of patients at the time of diagnosis confirmation. The second most common symptom was asthenia (4 out of 7; 57.1%), followed by dyspnea, cough and expectoration (3 out of 7; 42.9% for each symptom). Neurological (2 out of 7; 28.6%) and gastrointestinal (1 out of 7; 14.3%) symptoms were less common. Only 2 of the patients (28.6%) presented one single symptom from onset, while other 2 of cases presented two symptoms and 3 with three or more symptoms.

The MEWS score confirmed that Covid-19’s infection was severe in 5 out 7 of the patients (71.4%). A total of 3 patients required oxygen therapy, which was always applied at a low flow (less than 3l/minute) to achieve ≥98% oxygen saturation in the capillary oximetry continuous measure.

### Laboratory and radiological findings

COVID-19 infection was confirmed with RT-PCR in all the cases. Laboratory tests results and radiological results are shown in Table 2. Lymphocytopenia was found in 5 out of 7 of the patients (71.4%), but only one of them showed thrombocytopenia. Unfortunately, ferritin values were only available in 4 out of 7 of the cases, of which two presented normal values and the other two showed slightly elevated levels, below 1000 ng/ml. None of our patients showed altered levels of alanine aminotransferase, aspartate aminotransferase and LDH.

**TABLE 2.** Laboratory and radiological findings.

| **TABLE 2. Laboratory and radiological findings** |  |
| --- | --- |
| **Variable** | **Sample (N=7)** |
| White-cell count |  |
| <4000 per mm<sup>3</sup> – no./total no. (%) | 0/7 (0) |
| 4000-10.000 per mm<sup>3</sup> – no./total no. (%) | 6/7 (85.7) |
| >10.000 per mm<sup>3</sup> – no./total no. (%) | 1/7 (14.3) |
| Lymphocyte count |  |
| <1500 per mm<sup>3</sup> – no./total no. (%) | 5/7 (71.4) |
| Platelet count |  |
| <150.000 per mm<sup>3</sup> – no./total no. (%) | 1/7 (14.3) |
| Median hemoglobin (IQR) – g/dl | 11.1 (8.7-14.7) |
| Other findings– no./total no. (%) |  |
| C-reactive protein>10mg/dl | 7/7 (100) |
| Procalcitonin> 0.5ng/ml | 0/2 (0) |
| Lactatedehydrogenase>250U/liter | 0/7 (0) |
| Aspartateaminotransferase>40 U/liter | 0/7 (0) |
| Alanineaminotransferase> 40U/liter | 0/7 (0) |
| Total bilirubin>1mg/dl | 0/7 (0) |
| Urea > 40mg/dl | 0/7 (0) |
| Ferritin> 500ng/ml | 2/4 (50) |
| Radiologic findings |  |
| Abnormalities on chest radiograph – no./total no. (%) | 5/7 (71.4) |
| Ground-glass opacity – no./total no. (%) | 1/7 (14.3) |
| Unilobular pneumonia– no./total no. (%) | 1/7 (14.3) |
| Multilobular pneumonia – no./total no. (%) | 3/7 (42.9) |

With respect to the radiographic findings, 2 out of 7 of our patients showed no radiological alterations at diagnosis (Fig. 1A), 3 out of 7 of them presented bilateral multilobular pneumonia (Fig. 1B), 1 with unilobular pneumonia (Fig. 1C) and another one with a ground glass pattern (Fig. 1D). Only one case showed pleural effusion (Fig. 1C).

**Figure 1.**
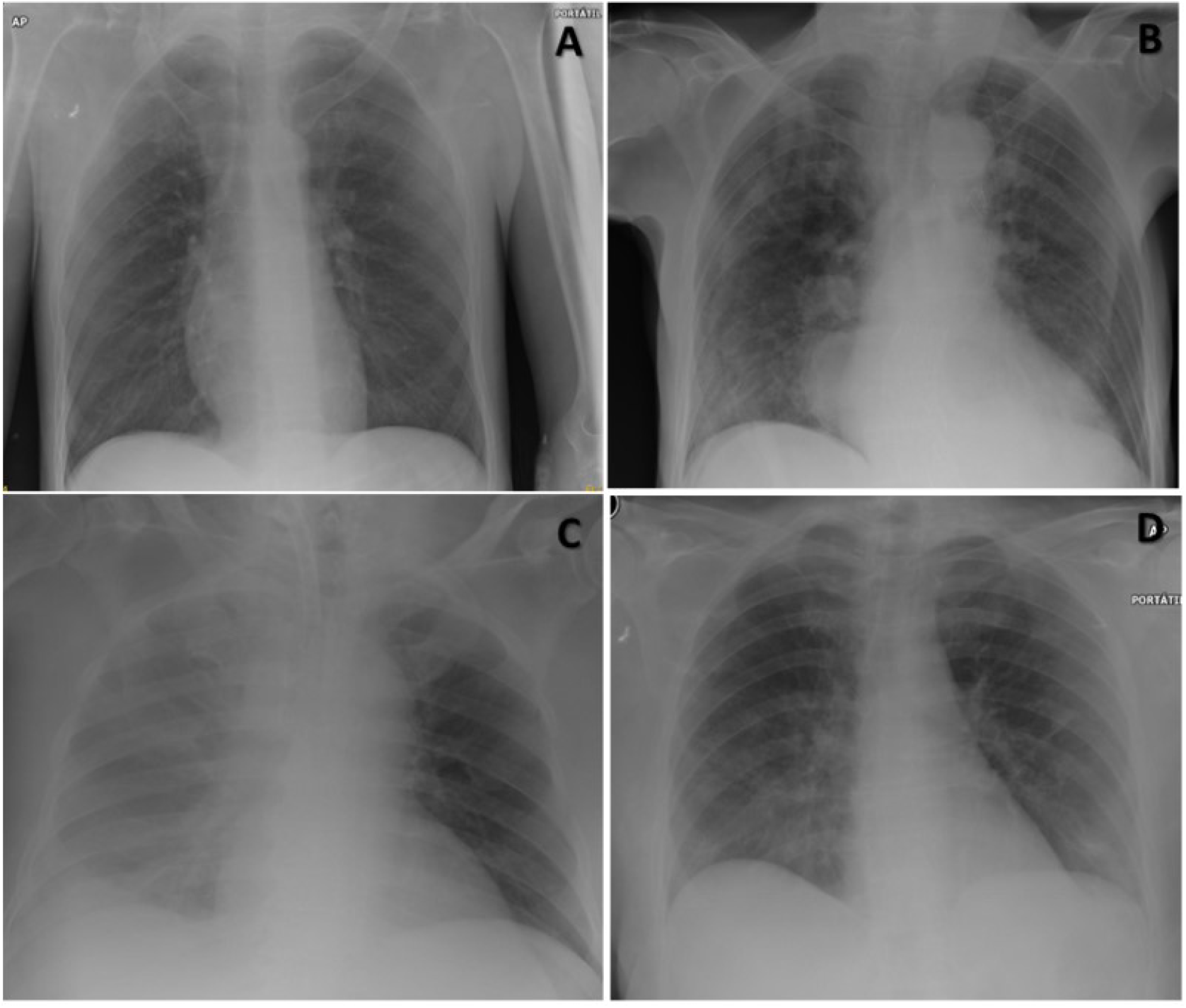
Representative radiological findings. **A)** Normal x-ray. **B)** Bilateral multiple lobular subsegmental pneumonia. **C)** Unilobular pneumonia with pleural effusion. **D)** Ground glass pattern.

### Clinical evolution

All of the patients have been gradually recovering, although the evolution was slower in the older patients, which were also depended on tracheostomy and suffered a cervical AIS A SCI. No deaths were recorded.

## Discussion

To the author’s knowledge, this is the first work that describes the clinical profile of Covid-19 in a cohort of SCI patients. The vulnerability of people with SCI is well known^6-11^. We have found that more than half of our patients showed one or two symptoms at the moment of Covid-19 confirmation, which is lower than the symptomatic expression in other published case series^18^. We hypothesized that the Covid-19 symptoms overlapped with some of the signs of respiratory failure due to SCI, which would have complicated the accurate clinical diagnosis. The average time delay between diagnosis in our case series was shorter than that found in other series, which may reflect rapid early identification of the initial Covid-19 symptoms in a cohort of hospitalized patients who were already receiving close attention for their SCI. At the onset of Covid-19 infection, fever and asthenia were the most frequently observed symptoms, which slightly differed from other published series that identified cough and fever as initial symptoms in the general population^18-20^. We agree with the previous case Covid 19 SCI in a patient with SCI which reported that cough was not the prevalent symptom^13^. However, despite the less evident symptomatology in our cohort, the MEWS score confirmed that they presented a higher risk of clinical worsening compared to the general population^18^. Before the clinical onset of Covid-19 infection, only half of the patients had a tracheostomy which required bronchial aspiration and hyperinflation techniques to remove respiratory secretions. In contrast, after Covid-19 diagnosis, all of the patients required such techniques. In other studies a similar progression in the requirement for respiratory assistance has also been observed for patients who require oxygen therapy^18-20^ with 98% capillary oxygen saturation achieved with low flow oxygen therapy (less than 3 liters/minute). To date, none of the patients with SCI and Covid-19 infection have required mechanical ventilation support or intensive care admission.

In agreement with most of the published studies, all of the COVID-19 cases were confirmed with RT-PCR. The radiological findings also agreed with the published data for Covid-19 infection, with the presentation of bilateral multilobe pneumonia as the most frequent radiological pattern. In addition, two thirds of our cases also presented lymphocytopenia^4,18,20^. However, we did not find any alteration of transaminases or LDH levels. Serum ferritin values were obtained in almost all the patients, with only half of them showing moderately elevated levels (< 1000 ng / ml). These results may highlight a decreased inflammatory response in our series compared to the general population^5,20^ and could be consistent a more benign evolution clinical evolution of Covid-19 infection with people with SCI.

We consider it of vital importance to closely supervise the SCI population to identify early compatible Covid-19 symptoms and signs, as well as to implement follow up measures against infectious diseases contagion. These measures will be especially needed in long-term treatment facilities such as our hospital, where asymptomatic cases could become Covid-19 reservoirs, further complicating the eradication of the current epidemic or future outbreaks. The limitations of this study include the fact that none of the patients admitted to the HNP or their healthcare personnel were interviewed for symptoms of infection or received the SARS-CoV-2 screening test, which therefore may have led to an underestimation of the Covid-19 infection rate in the hospital, especially in those patients who were asymptomatic or who presented mild symptoms.

As this is a descriptive case series study, there is no control group, the absence of which is justified given the urgency of pandemic and the lack of previous Covid-19 data among SCI population. Unfortunately, it is likely that our Covid-19 SCI case series will gradually increase until the end of the present pandemic, which will provide us with more clinical data with longer evolution times.

## Conclusion

In our case series, patients with SCI and confirmed Covid-19 infection exhibited fewer symptoms than the general population. Furthermore, although they presented a similar or greater MEWS severity the clinical evolution of Covid-19 infection was not as pronounced as had been expected. This study recommends close active supervision of the SCI population to detect early compatible signs and symptoms of Covid-19 infection.

## Data Availability

The main sources of data were the electronic medical records and clinical reports of each one of the patients. The history of recent contact with a confirmed case, the diagnosis time delay, the symptom and signs of onset and evolution, the laboratory and chest X-rays were analyzed. In order to avoid biases and ensure the patients' data confidentiality, all documents were handled after removing their filiation data. The corresponding author had full access to all data in the study and had final responsibility for the decision to submit for publication.

## Acknowledgements

Dr. Julian Taylor (JTG) revised the manuscript and reviewed the English version.

## Statement of ethics

We certify that all applicable institutional and governmental regulations concerning the ethical use of human volunteers were followed during the course of this research. All participants provided their informed consent. The guidelines of the declaration of Helsinki were followed in every case and the study design was approved by the local ethics committee.

## Author contribution

MRC was responsible for designing the clinical protocol and collecting clinical and analytical data, conducting the research, analysing data, interpreting results and writing the manuscript. IJV was responsible for designing the clinical protocol and collecting clinical and analytical data. FGH was responsible for designing the clinical protocol and collecting clinical and analytical data.

ELD was responsible for designing the clinical protocol and collecting clinical and analytical data, identifying bibliographic resources, analysing data, interpreting results and writing the manuscript

CGM was responsible for designing the clinical protocol and conducting the research.

EVB was responsible for designing the clinical protocol and collecting clinical and analytical data.

JTG was responsible for content revision and correction of English version

AGA was responsible for designing the clinical protocol, reviewing data quality and, conducting the research, analysing data, interpreting results and writing the manuscript.

## REFERENCES

1. Qun Li, Xuhua Guan, Peng Wu, et al. Early Transmission Dynamics in Wuhan, China, of Novel Coronavirus–Infected Pneumonia. NEJM 2020, January 29. DOI: 10.1056/NEJMoa2001316.

2. Coronavirus COVID-19 Global Cases by the Center for Systems Science and Engineering (CSSE) at Johns Hopkins University (JHU) [Internet]. JHU COVID-19 Resource Center. Johns Hopkins Coronavirus Resource Center; [Accessed 2020 April 2]: https://www.arcgis.com/apps/opsdashboard/index.html#/bda7594740fd40299423467b48e9ecf6.

3. Protocolo de actuación frente a casos de infección por el Nuevo Coronavirus (SARS-CoV-2). Ministerio de Sanidad. Gobierno de España. Accesed April 4, 2020. https://www.mscbs.gob.es/profesionales/saludPublica/ccayes/alertasActual/nCovChina/documentos/Procedimiento_COVID_19.pdf

4. Zhou F et al. Clinical course and risk factors for mortality of adult inpatients with COVID-19 in Wuhan, China: a retrospective cohort study.Lancet. 2020 Mar 11. pii: S0140-6736(20)30566-3. doi: 10.1016/S0140-6736(20)30566-3.

5. Xin Sun et al. Multiple organ dysfunction and systemic inflammation after spinal cord injury: a complex relationship. Journal of Neuroinflammation (2016) 13:260. DOI 10.1186/s12974-016-0736-y.

6. Riegger, T. et al. Spinal cord injury-induced immune depression syndrome (SCI-IDS). Eur. J. Neurosci 2007, 25:1743–1747.

7. Kasinathan N, Vanathi MB, Subrahmanyam VM, Rao JV. A Review on Response of Immune System in Spinal Cord Injury and Therapeutic Agents useful in Treatment. CurrentPharmaceuticalBiotechnology, 2015 (16):26–34.

8. Prüss H, Tedeschi A, Thiriot A, Lynch L, Loughhead SM, Stutte S,^7^, Mazo IB, Kopp MA, Brommer B, Blex C, Geurtz LC, Liebscher T, Niedeggen A, Dirnagl U, Bradke F, Volz MS, DeVivo MJ, Chen Y, von Andrian UH, Schwab JM. Spinal cord injury-induced immunodeficiency is mediated by a sympathetic-neuroendocrine adrenal reflex.NatNeurosci. 2017 Nov;20(11):1549–1559. doi: 10.1038/nn.4643.

9. Arévalo-Martin A, et al. Elevated Autoantibodies in Subacute Human Spinal Cord Injury Are Naturally Occurring Antibodies. Front. Immunol. (2018) 9:2365. doi: 10.3389/fimmu.2018.02365

10. Shah A, Shem K, McKENNA S, Berlly M. Respiratory management of the spinal cord injured patients. In: Kirshblum S, Campagnolo DI. Spinal Cord Medicine. 2011. Lippincott Williams & Wilkins, Philadelphia.

11. Kopp, M.A. et al. Long-term functional outcome in patients with acquired infections after acute spinal cord injury. Neurology 2017, (88):892–900.

12. Stillman MD, Capron M, Alexander M, Longini Di Giusto M, Scivoletto G. COVID-19 and spinal cord injury and disease: results of an international survey. Spinal Cord Series and Cases 2020,6:21

13. Righi G, Del Popolo G. COVID-19 tsunami: the first case of a spinal cord injury patient in Italy. Spinal Cord Series and Cases 2020,6:22

14. American Injury Association: International Standards of Neurological Classification of Spinal Cord Injury, revised 2019; Richmond VA.

15. Zu ZY, Jiang MD, Xu PP, Chen W, Ni QQ, Lu GM, Zhang LJ. Coronavirus Disease 2019 (COVID-19): A Perspective from China.Radiology, 2020 Feb 21:200490. doi: 10.1148/radiol.2020200490.

16. Subbe C, Kruger M, Rutherford P, Gemmel L. Validation of a modified Early Warning Score in medical admissions. Qjm.2001;94(10):521–6.

17. Lim WS, van der Eerden MM, Laing R, Boersma WG, Karalus N, Town GI, et al. De?ning community acquired pneumonia severity on presentation to hospital: an international derivation and validation study. Thorax 2003, 58:377–382.

18. Guan WJ et al. China Medical Treatment Expert Group for Covid-19. Clinical Characteristics of Coronavirus Disease 2019 inChina.NEngl J Med. 2020 Feb 28th. doi: 10.1056/NEJMoa2002032

19. Fauci AS. Covid-19—Navigating the Uncharted. N Engl J Med 2020; 382:1268–1269 DOI: 10.1056/NEJMe2002387

20. Huang C, et al.Clinical features of patients infected with 2019 novel coronavirus in Wuhan, China. Lancet 2020; 395: 497–506.

